## Supplementary File for "Examining causal pathways between family adversity and soiling: a prospective cohort study"

### Supplementary Text

#### Methods: Sample

ALSPAC is a prospective, population-based birth cohort study that recruited pregnant women resident in Avon, UK with expected dates of delivery between 1st April 1991 and 31st December 1992 [1,2]. The initial number of pregnancies enrolled was 14,541 resulting in 14,062 live births and 13,988 children who were alive at 1 year of age. When the oldest children were approximately 7 years of age, an attempt was made to bolster the initial sample with eligible cases who had failed to join the study originally, resulting in an additional 913 children being enrolled. The total sample size for analyses using any data collected after the age of seven is therefore 15 447 pregnancies, resulting in 15,658 foetuses. Of these 14,901 children were alive at 1 year of age. 14,203 unique mothers were initially enrolled in the study and 12,113 partners have been in contact with the study of which 3,807 are enrolled. The phases of enrolment are described in more detail in the cohort profile paper and its update [1,2]. The study website ([www.bristol.ac.uk/alspac](http://www.bristol.ac.uk/alspac)) contains details of all the data that is available through a fully searchable data dictionary and variable search tool:	<http://www.bristol.ac.uk/alspac/researchers/our-data/>. Ethical approval for ALSPAC was obtained from the ALSPAC Ethics and Law Committee and the Local Research Ethics Committees. Informed consent for the use of data collected via questionnaires and clinics was obtained from participants following the recommendations of the ALSPAC Ethics and Law Committee at the time.

#### Methods: The Family Adversity Index (FAI)

The FAI was derived from responses to questions asked about ALSPAC participants (parents and children) that reflected the following family-based risk factors:

- **Age of mother^*^** (1): at first pregnancy/child’s birth
- **Housing** (3): a) adequacy, b) basic living, c) defects/infestation
- **No educational qualifications^*^** (1) achievement (mother or father)
- **Financial status** (1): a) financial difficulties
- **Partner relationship** (4): a) status, b) affection and aggression, c) physical/emotional cruelty, d) no social support
- **Family** (2): a) family size, b) Major problems (child in care, not with natural mother, or on Social Services at-risk register^**^)
- **Social network** (2): a) emotional support, b) Practical/financial support
- **Maternal Psychopathology** (1): a) Affective (depression, anxiety, or suicide attempts)
- **Substance abuse** (1): a) drugs or alcohol (use of hard drugs, alcoholism, alcohol consumption)
- **Crime** (2): a) In trouble with police, or b) convictions

*[The numbers in brackets refer to the number of items within each category. *Note that these items were measured during the antenatal period or at birth. **The at-risk register has been superseded by the Child Protection Plan (CPP); The Children Act 1989]*

The FAI scores were calculated where >50% of the above items were valid and non-adversity was assumed for any missing data. Each item was assigned a value of 1 if the adversity was present and 0 if not. The final score was derived from the sum of items giving a possible score range from 0 to 18 and the higher the score recorded the greater the level of adversity.

Details of the questionnaires from which these items were derived are available in the ALSPAC study documentation file D4297_mLon.pdf held in the publicly available data dictionary: https://www.bristol.ac.uk/alspac/external/documents/ALSPAC_Data_Dictionary.zip

#### Methods: The parametric g-formula

In mediation analysis, four assumptions are made with respect to confounding: no unmeasured confounders for the exposure-outcome, exposure-mediator or mediator-outcome paths; no measured or unmeasured confounders of the mediator-outcome path that lie on the causal pathway from the exposure. We implemented the parametric g-computation approach, which relaxes the assumption of no measured intermediate confounders [3,4].

We used the Stata package *gformula* to conduct parametric g-computation to estimate the total causal effect (TCE), natural direct effect (NDE) and the natural indirect effect (NIE) [3]. G-formula uses Monte Carlo simulations to simulate the outcome, mediators and intermediate confounders under hypothetical interventions or “counter to the fact” scenarios (i.e. contrary to the intervention individuals actually received). The TCE is the value the outcome (e.g. daytime soiling) would take if all individuals had been exposed to a one-unit increase in the FAI versus everyone not being exposed to a one unit increase in the FAI. The NDE is the direct (unmediated) effect of the exposure (e.g. FAI) on the outcome (e.g. daytime soiling) when the mediator(s) (e.g. constipation) takes the value it would be in absence of the exposure (i.e. FAI of 0). More specifically it is modelled as the direct effect of exposure X = x+1 (e.g. an FAI of 1) versus exposure X = x (e.g. an FAI of 0) on outcome Y (e.g. daytime soiling) if mediator M (e.g. constipation) were set to whatever value it would be for X = x. The NIE is the effect of the exposure (e.g. FAI) on the outcome (e.g. daytime soiling) that operates by changing the mediator (e.g. constipation). More specifically, it is modelled as the effect on outcome Y (e.g. daytime soiling) if the exposure X = x+1 and the mediator M (e.g. constipation) were changed from the value it would take if X = x to the value it would take if X = x+1. The proportion of the TCE that is mediated is calculated as [OR_NDE_ (OR_NIE_ − 1)] / [OR_NDE_ × OR_NIE_ − 1] × 100 [5].

Specifically for this analysis we used a Monte Carlo sample size of 100,000 to minimise fluctuations in effect estimates. Standard errors were estimated using 250 bootstrap samples. We did not hypothesise an interaction between the exposure and mediators and therefore did not include an interaction term within the model. We also specified the command option *linexp* to specify that the exposure is continuous, and its effect is assumed to be linear. For the imputed data we conducted mediation analysis for each of the 50 imputation datasets and output the results to log files. We used R to extract the log ORs and bootstrapped SEs for each of the TCE, NDE, and NIE from each Stata log file. The mean of the log OR was computed and the SEs were calculated using Rubin’s rules [6]. The 95% confidence intervals were calculated from the combined log OR and SEs. Log ORs (and 95% CIs) were exponentiated to represent ORs.

#### Methods: Missing data

We used multivariate imputation by chained equations under the missing at random assumption to impute missing data [7] in daytime soiling and baseline and intermediate confounders up to the sample with complete data on the FAI score (N = 10,033). We included two types of auxiliary variables; i) additional measures (both before and/or after) of those of the substantive model, and ii) sociodemographic indicators associated with both missingness and many of the variables of the substantive model. These serve two main functions: 1) increasing precision by improving the predictive ability of the imputation model beyond that of the substantive model [8] and 2) making the MAR assumption more plausible by reducing the bias due to data being MNAR [9] (see Supplementary Table S2). All binary variables were imputed using logistic regression, categorical (unordered) variables were imputed using polytomous regression, and all continuous variables were imputed using predictive mean matching. Missing data were imputed separately in males and females before appending the two datasets together. For both males and females 50 imputed datasets with 50 iterations were created using the mice package (version 3.16.0) in R. Estimates were then combined using Rubin’s rules [6].

#### Results: Description of sample

Table 1 in the main manuscript shows the distribution of study variables in the imputed (N = 10 033) and complete case samples (N = 3366). Any daytime soiling was reported for 7.3% of children. In total, 71.7% of children experienced one or more family adversities, the most prevalent being maternal affective disorders (depression, anxiety, and/or suicide attempt; 20.5%). A lack of social support from partner relationships was also common (19.6%; see Supplementary Table 3 for all component prevalences). The mean number of family adversities experienced was 1.9 (Standard Error (SE) 0.02) in all children and was higher in those experiencing soiling (2.3, SE 0.10) compared to those without soiling (1.8, SE 0.02; ttest p-value <0.001). Compared to the imputed data sample, children in the complete case sample experienced fewer adversities and lower proportions of non-white ethnicity, manual social class, preterm birth, parental abuse, and temper tantrums. The complete case sample also had a higher proportion of children with picky eating, hard stools, no maternal smoking during pregnancy, and maternal married status.

### Supplementary Tables

#### Table S1 Variable description and coding

| **Variable name** | **ALSPAC variable^1^** | **Time of measure** | **Measure/Question** | **Responses** | **Final variable coding** |
| --- | --- | --- | --- | --- | --- |
| ***Exposure*** | | | | | |
| **Family Adversity Index (FAI)**  From 0-2 years | blong | 0 to 2 years | DV: long index comprising of 18 items including information on the age of the mother, housing, education, financial status, partner relationship, family, social network, maternal psychopathology, substance abuse and crime | Continuous score (0-18) | Continuous score (0-18) |
| ***Outcome*** | | | | | |
| **Daytime soiling** Frequency child dirties pants during the day | kr125 | 7.6 years | How often usually does your child dirty his/her pants during the day? | 1: Never 2: < once /week 3: ~ once / week 4: 2-5 times / week 5: Nearly every day 6: > once / day -6; -1: Missing | Recode: 1=0: No 2/6=1: Yes |
| ***Mediators*** | | | | | |
| **Emotional/behaviour problems** Parent version of the Strengths and Difficulties Questionnaire by Goodman (1997) Total difficulties score | j557f | 3.9 years | Derived variable: prorated score of the first 4 subscales (hyperactivity, conduct problems, emotional problems, and peer problems) | Continuous; missing = -1 | Continuous (score 0-33) |
| **Constipation** | kn1037 | 5.8 years | In the past 15 months child has had constipation? | 1: Yes & Saw Doctor 2: Yes But Did Not See Doctor 3: Did Not Have -11; -10; -1: Missing | Recode: 3=0: No 1/2=1: Yes |
| ***Baseline Confounders*** | | | | | |
| **Ethnicity**  Child ethnic background | c804 | Antenatal | Derived variable: Ethnic group reported for the mother (respondent) or partner: Non-white if c801 or c802 had codes in the range of 2-9 | 1: White 2: Non-white -1: Missing | Recode: 1=0: White 2=1: Non-white |
| **Sex** Child’s sex assigned at birth | kz021 | Birth | Recorded at birth by the fieldworkers who visited the maternity units | 1: Male 2: Female -2; -1: Missing | Recode: 1=0: Male 2=1: Female |
| **Preterm birth** Term vs. preterm | bestgest | Birth | Derived variable: Best guess of gestation when pregnancy ended | Continuous (range 24-47) -11; -10; -3; -2: Missing | Recode: >36 weeks=0: Term ≤36 weeks=1: Preterm |
| **Marital status** | a525 | Antenatal | What is your present marital status? | 1: Never married 2: Widowed 3: Divorced 4: Separated 5: 1st marriage 6: Marriage 2 or 3 -1: Missing | Recode: 5/6=0: Married 1=1: Single 2/4=2: Divorced/separated/widowed |
| **Smoking** Maternal smoking during pregnancy | b665 (1st trimester) | Antenatal | Did you smoke regularly in the first three months of pregnancy? | 1: No 2: Yes, cigarettes 3: Yes, cigars 5: Yes, other | Recode: [No (0) smoking during trimesters 1, 2 and 3]=0: None [Smoked during 1st trimester (2, 3, 5) but not the 2nd (0) and 3rd (0)]=1: Yes, quit early [Smoked during trimesters 1 (2, 3, 5), 2 (2, 3, 5), and 3 (1, 2, 3) or either 1 and 2 or 1 and 3, or 2 and 3]=2: Yes, throughout |
|  | b667 (2nd trimester) | Antenatal | Did you smoke regularly in the last 2 weeks? | 1: No 2: Yes, cigarettes 3: Yes, cigars 5: Yes, other |  |
|  | c483 (3rd trimester) | Antenatal | How many cigarettes per day are you yourself smoking at the moment? | 0: None 1: 1-9 2: 10-19 3: 20+ -7: Missing |  |
| **Household social class** 1991 British Office of Population and Census Statistics (OPCS) job codes (maternal) 1991 British Office of Population and Census Statistics (OPCS) job codes (paternal) | c755  c765 | Antenatal | Derived variable from questions:  *Actual job, occupation, trade or profession   *Please tick which of the following apply to you: foreman, manager, supervisor, leading hand, self-employed, none of these *Type of industry or service given (main things done in job) | 1 - I  2 - II 3 – III (non-manual)  4 – III (manual)  5 - IV  6 - V  65 - Armed forces  -1 - missing | Highest parental social class i.e. if one parent is non-manual then social class=0:   [Professional, managerial or skilled professions (responses 1, 2, 3)]=0: Non-manual [Partly or unskilled occupations (responses 4, 5, 6)]=1: Manual  65=missing |
| ***Intermediate Confounders*** | | | | | |
| **Body mass index (BMI)** | bmi_chdb4 |  | Child health database measures of BMI: Derived from measure of height (m) and weight (kg). Calculated as kg/m2 | Continuous measure | Continuous measure |
| **Difficult temperament** Emotionality, Activity, and Sociability (EAS) Scale | kg620b | 3.2 years | Derived variable: EAS emotionality subscale (prorated) | Continuous (score 5-25) -6; -5: Missing | Continuous (score 5-25) |
| **Temper tantrums** Frequency child has temper tantrums | kj255 | 3.5 years | How often does he/she have temper tantrums? | 1: Once per Day 2: Most Days 3: > Once a WK 4: < Once a WK 5: Never -1: Missing | Recode: 3/4/5=0: Few or no temper tantrums 1/2=1: Once a day or most days |
| **Developmental delay** Total developmental score | kf607 | 2.5 years | Derived variable: prorated scores | Continuous (age-adjusted z-score); missing = -102; -101 | Continuous (z score) |
| **Hard stools** Frequency child's stools are hard | kf141 | 2.5 years | Nowadays how often are his/her stools hard? | 1: Usually 2. Sometimes 3. Never -1: Missing | Recode: 2/3=0: No 1=1: Yes |
| **Stool frequency** Number of motions child has everyday | kf140 | 2.5 years | Nowadays how many motions (or dirty nappies) a day (24 hours) does he usually have? | 0: Other 1: 4 or > 4 times 2: 2 - 3 times 3: once a day 4: once in 2 - 4 days 5: once a week 9: can't say -1: missing | Recode: 1/2/3=0: Frequent 4/5=1: Infrequent 9=missing |
| **Picky eating** Likes and dislikes of food | kg466 | 3.2 years | Does your child have definite likes and dislikes as far as food is concerned? | 1: No eats most food 2. Yes quite choosy 3. Yes very choosy -1: Missing | Recode: 1/2=0: No 3 =1: Yes, picky eater |
| **Diet: Fibre intake** PC: Healthy eating (proxy for fibre) Northstone et al 2000 | kg521 | 3.2 years | Derived variable: PCA score 2 'healthy' 38 months | Continuous | Continuous |
| **Maternal bond** Maternal bonding score | h766 | 2.8 years | Derived variable from h760 + h763 | Continuous score (0-33) -1: Missing | Continuous score (0-33) |
| **Parental abuse** Parental cruelty (physical and emotional) Question: Have any of these occurred since the study child was 18 months old? | h236a | 2.8 years | Partner physically cruel to children | 1: Yes 2: No -1: Missing | Recode: [h236a, h237a, h247a, h248a: 2]=0: No [any h236a, h237a, h247a, h248a: 1]=1: Yes For imputation derive: Physical cruelty [h236a, h237a: 2]=0: No / [h236a, h237a: 1]=1: Yes Emotional cruelty [h247a, h248a: 2]=0: No / [h247a, h248a: 1]=1: Yes Then passively impute:  Physical cruelty = No & Emotional cruelty = No ==0: No parental abuse Physical cruelty = Yes &/or Emotional cruelty = Yes ==1: Yes parental abuse |
|  | h237a | 2.8 years | Mother physically cruel to children | 1: Yes 2: No -1: Missing |  |
|  | h247a | 2.8 years | Partner emotionally cruel to children | 1: Yes 2: No -1: Missing |  |
|  | h248a | 2.8 years | Mother emotionally cruel to children | 1: Yes 2: No -1: Missing |  |

^1^ ALSPAC variable names; these can be searched for using the ALSPAC variable search tool [http://variables.alspac.bris.ac.uk/].

#### Table S2 Auxiliary variables for multiple imputation of missing data

Daytime soiling in child measured at age 6

Daytime soiling in child measured at age 6

Total difficulties score measured at age 6

Constipation in child measured at age 8

Stool frequency measured at age 3

Hard stools measured at age 3

BMI measured at age 18 months

Picky eating measured at age 2

Healthy eating measured at age 2

House tenure measured in antenatal period

Household crowding measured in antenatal period

Material hardship measured in antenatal period

Antenatal depression in mothers measured in antenatal period

Antenatal anxiety in mothers measured in antenatal period

#### Table S3 Prevalences of the individual adversity components of the family adversity index in (N = 10,033)

| FAI constituent | Sub-measure affective group | Prevalence (%) |
| --- | --- | --- |
| Age of mother | Early parenthood | 5.2 |
| Crime | Been in trouble with police | 3.7 |
| Crime | Has convictions | 0.6 |
| Family | High family size | 5.5 |
| Family | Has family major problems | 0.9 |
| Financial status | financial difficulties | 15.3 |
| Housing | Has housing defects/infestation | 17.5 |
| Housing | Poor housing adequacy | 8.6 |
| Housing | Poor housing - basic living | 4.8 |
| Maternal psychopathology | Has affective disorders (depression, anxiety and/or suicide attempts) | 20.5 |
| No educational qualifications | Low educational achievement (mother or father) | 12.4 |
| Partner relationship | Lack of partner social support | 19.6 |
| Partner relationship | Lack of partner affection | 14.7 |
| Partner relationship | Experienced partner cruelty | 12.9 |
| Partner relationship | Partner relationship status | 6.5 |
| Social network | Lack of emotional support | 12.6 |
| Social network | Lack of practical/financial support | 11.2 |
| Substance abuse | Substance/alcohol abuse | 13.4 |

FAI Family adversity index

#### Table S4 Results for the associations between family adversity index and mediators in the imputed data sample (N = 10,033)

| **Exposure: FAI** | | **Univariable** | | | **Multivariable** | | |
| --- | --- | --- | --- | --- | --- | --- | --- |
|  |  | **Beta/OR** | **95% CI** | **p-value** | **Beta/OR** | **95% CI** | **p-value** |
| **Outcome** | Emotional/behaviour problems | 0.587 | 0.537, 0.638 | <0.001 | 0.524 | 0.471, 0.577 | <0.001 |
|  | Constipation | 1.086 | 1.047, 1.125 | <0.001 | 1.072 | 1.033, 1.113 | <0.001 |

OR odds ratio; CI confidence interval; FAI family adversity index; all multivariable models adjusted for baseline confounders (sex, ethnicity, preterm birth, household social class, marital status, and smoking during pregnancy). Parameter estimates for emotional/behaviour problems are Betas; constipation are ORs.

#### Table S5 Results for the associations between emotional/behaviour problems and constipation in the imputed data sample (N = 10,033)

| **Exposure: Emotional/behaviour problems** | **Univariable** | | | **Multivariable** | | |
| --- | --- | --- | --- | --- | --- | --- |
|  | **OR** | **95% CI** | **p-value** | **OR** | **95% CI** | **p-value** |
| Constipation | 1.033 | 1.017, 1.050 | <0.001 | 1.007 | 0.986, 1.027 | 0.521 |

OR odds ratio; CI confidence interval; multivariable models adjusted for baseline confounders (sex, ethnicity, preterm birth, household social class, marital status, and smoking during pregnancy) and the Family Adversity Index.

#### Table S6 Results for the associations between family adversity index and mediators with daytime soiling in the complete case sample (N = 3,366)

| **Outcome: daytime soiling** | | **Univariable** | | | **Multivariable** | | |
| --- | --- | --- | --- | --- | --- | --- | --- |
|  |  | **OR** | **95% CI** | **p-value** | **OR** | **95% CI** | **p-value** |
| **Exposure** | FAI | 1.080 | 0.998, 1.165 | 0.051 | 1.104 | 1.015, 1.196 | 0.019 |
|  | Emotional/behaviour problems^1^ | 1.029 | 0.999, 1.061 | 0.059 | 0.995 | 0.959, 1.032 | 0.805 |
|  | Constipation^1,2^ | 3.309 | 2.355, 4.591 | <0.001 | 3.596 | 2.525, 5.063 | <0.001 |

OR odds ratio; CI confidence interval; FAI family adversity index; all multivariable models adjusted for baseline confounders (sex, ethnicity, preterm birth, household social class, marital status, and smoking during pregnancy). ^1^ Models additionally adjusted for intermediate confounders (child developmental delay, maternal bonding score, parental abuse, stool frequency, hard stools, difficult temperament, picky eating, healthy eating, temper tantrums, and body mass index), and FAI. ^2^ Model additionally adjusted for emotional/behaviour problems. Parameter estimates are based on a one-unit increase in FAI, a one-unit increase in emotional/behaviour problems score, and the comparison between yes vs. no for constipation.

#### Table S7 Results for the associations between family adversity index and mediators in the complete case sample (N = 3,366)

| **Exposure: FAI** | | **Univariable** | | | **Multivariable** | | |
| --- | --- | --- | --- | --- | --- | --- | --- |
|  |  | **Beta/OR** | **95% CI** | **p-value** | **Beta/OR** | **95% CI** | **p-value** |
| **Outcome** | Emotional/behaviour problems | 0.625 | 0.534, 0.717 | <0.001 | 0.585 | 0.490, 0.680 | <0.001 |
|  | Constipation | 1.100 | 1.031, 1.171 | 0.003 | 1.086 | 1.015, 1.160 | 0.015 |

OR odds ratio; CI confidence interval; FAI family adversity index; all multivariable models adjusted for baseline confounders (sex, ethnicity, preterm birth, household social class, marital status, and smoking during pregnancy). Parameter estimates for emotional/behaviour problems are Betas; constipation are ORs.

#### Table S8 Results for the mediation of family adversity index and daytime soiling via emotional/behaviour problems and constipation in the complete case sample (N = 3,366)

| **Total causal effect** | | **Natural indirect effect** | | **Natural direct effect** | | **PM%** |
| --- | --- | --- | --- | --- | --- | --- |
| **OR** | **95% CI** | **OR** | **95% CI** | **OR** | **95% CI** |  |
| 1.092 | 1.007, 1.215 | 0.997 | 0.957, 1.017 | 1.096 | 1.023, 1.017 | NA |

OR odds ratio; CI confidence interval; PM proportion mediated (%). Mediation models were fitted with emotional/behaviour problems and constipation as the mediators of interest; child developmental delay, maternal bonding score, parental abuse, stool frequency, hard stools, difficult temperament, picky eating, healthy eating, temper tantrums, and body mass index were considered intermediate confounders. All paths were adjusted for baseline confounders: sex, ethnicity, preterm birth, household social class, and smoking during pregnancy.

### Strobe statement

**STROBE Statement—checklist of items that should be included in reports of observational studies**

|  | Item No | Recommendation |
| --- | --- | --- |
| **Title and abstract**  **See title and abstract** | 1 | (*a*) Indicate the study’s design with a commonly used term in the title or the abstract |
|  |  | (*b*) Provide in the abstract an informative and balanced summary of what was done and what was found |
| Introduction | | |
| Background/rationale  **Introduction paragraphs 1 & 2** | 2 | Explain the scientific background and rationale for the investigation being reported |
| Objectives  **Introduction paragraph 3, Figure 2** | 3 | State specific objectives, including any prespecified hypotheses |
| Methods | | |
| Study design  **Methods** | 4 | Present key elements of study design early in the paper |
| Setting  **Methods: Sample, Supplementary Text** | 5 | Describe the setting, locations, and relevant dates, including periods of recruitment, exposure, follow-up, and data collection |
| Participants  **Methods: sample, Figure 1, Supplementary Text** | 6 | (*a*) *Cohort study*—Give the eligibility criteria, and the sources and methods of selection of participants. Describe methods of follow-up  *Case-control study*—Give the eligibility criteria, and the sources and methods of case ascertainment and control selection. Give the rationale for the choice of cases and controls  *Cross-sectional study*—Give the eligibility criteria, and the sources and methods of selection of participants |
|  |  | (*b*) *Cohort study*—For matched studies, give matching criteria and number of exposed and unexposed  *Case-control study*—For matched studies, give matching criteria and the number of controls per case |
| Variables  **Methods, Supplementary Tables S1 & S2** | 7 | Clearly define all outcomes, exposures, predictors, potential confounders, and effect modifiers. Give diagnostic criteria, if applicable |
| Data sources/ measurement  **Methods, Supplementary Tables S1, Supplementary Text** | 8* | For each variable of interest, give sources of data and details of methods of assessment (measurement). Describe comparability of assessment methods if there is more than one group |
| Bias  **Methods: Statistical analysis and missing data, Supplementary Text, Discussion** | 9 | Describe any efforts to address potential sources of bias |
| Study size  **Methods: Sample, Figure 1, Supplementary Text** | 10 | Explain how the study size was arrived at |
| Quantitative variables  **Methods: Statistical analysis, Supplementary Table S1** | 11 | Explain how quantitative variables were handled in the analyses. If applicable, describe which groupings were chosen and why |
| Statistical methods  **Methods: Statistical analysis, Supplementary Text** | 12 | (*a*) Describe all statistical methods, including those used to control for confounding |
|  |  | (*b*) Describe any methods used to examine subgroups and interactions |
|  |  | (*c*) Explain how missing data were addressed |
|  |  | (*d*) *Cohort study*—If applicable, explain how loss to follow-up was addressed  *Case-control study*—If applicable, explain how matching of cases and controls was addressed  *Cross-sectional study*—If applicable, describe analytical methods taking account of sampling strategy |
|  |  | (*e*) Describe any sensitivity analyses |
| Results | | |
| Participants  **Figure 1, Supplementary Text, Table 1** | 13* | (a) Report numbers of individuals at each stage of study—eg numbers potentially eligible, examined for eligibility, confirmed eligible, included in the study, completing follow-up, and analysed |
|  |  | (b) Give reasons for non-participation at each stage |
|  |  | (c) Consider use of a flow diagram |
| Descriptive data  **Table 1, Results** | 14* | (a) Give characteristics of study participants (eg demographic, clinical, social) and information on exposures and potential confounders |
|  |  | (b) Indicate number of participants with missing data for each variable of interest |
|  |  | (c) *Cohort study*—Summarise follow-up time (eg, average and total amount) |
| Outcome data  **Table 1, Results** | 15* | *Cohort study*—Report numbers of outcome events or summary measures over time |
|  |  | *Case-control study—*Report numbers in each exposure category, or summary measures of exposure |
|  |  | *Cross-sectional study—*Report numbers of outcome events or summary measures |
| Main results  **Results, Tables 1-3, Supplementary Tables S3-7** | 16 | (*a*) Give unadjusted estimates and, if applicable, confounder-adjusted estimates and their precision (eg, 95% confidence interval). Make clear which confounders were adjusted for and why they were included |
|  |  | (*b*) Report category boundaries when continuous variables were categorized |
|  |  | (*c*) If relevant, consider translating estimates of relative risk into absolute risk for a meaningful time period |
| Other analyses  **Complete case analysis Supplementary Tables S5-7** | 17 | Report other analyses done—eg analyses of subgroups and interactions, and sensitivity analyses |
| Discussion | | |
| Key results  **Discussion paragraph 1** | 18 | Summarise key results with reference to study objectives |
| Limitations  **Discussion: Strengths and limitations** | 19 | Discuss limitations of the study, taking into account sources of potential bias or imprecision. Discuss both direction and magnitude of any potential bias |
| Interpretation  **Discussion: Comparison with previous findings and potential mechanisms** | 20 | Give a cautious overall interpretation of results considering objectives, limitations, multiplicity of analyses, results from similar studies, and other relevant evidence |
| Generalisability  **Discussion: Strengths and limitations** | 21 | Discuss the generalisability (external validity) of the study results |
| Other information | | |
| Funding  **Funding section** | 22 | Give the source of funding and the role of the funders for the present study and, if applicable, for the original study on which the present article is based |

*Give information separately for cases and controls in case-control studies and, if applicable, for exposed and unexposed groups in cohort and cross-sectional studies.

**Note:** An Explanation and Elaboration article discusses each checklist item and gives methodological background and published examples of transparent reporting. The STROBE checklist is best used in conjunction with this article (freely available on the Web sites of PLoS Medicine at http://www.plosmedicine.org/, Annals of Internal Medicine at http://www.annals.org/, and Epidemiology at http://www.epidem.com/). Information on the STROBE Initiative is available at www.strobe-statement.org.
